# Disproportionately Elevated Sulcal Index (DESI): An automatically driven index representing disproportionate subarachnoid space enlargement in brain MRI scans

**DOI:** 10.64898/2025.12.01.25341388

**Authors:** Siavash Shirzadeh Barough, Shin Ohno, Murat Bilgel, Haris I. Sair, Mark G. Luciano, Abhay Moghekar

## Abstract

**Background:** Idiopathic normal pressure hydrocephalus (iNPH) is a reversible cause of dementia in older adults yet remains underdiagnosed due to nonspecific clinical presentation and reliance on invasive confirmatory tests. Conventional MRI biomarkers, including the Evans index and callosal angle, capture ventricular enlargement but incompletely characterize cerebrospinal fluid (CSF) redistribution. Disproportionately enlarged subarachnoid-space hydrocephalus (DESH) features, tight high-convexity sulci and enlarged Sylvian fissures, are more disease-specific but lack standardized quantitative measures.

**Methods:** We developed the Disproportionately Elevated Sulcal Index (DESI), an automated volumetric biomarker quantifying the ratio of Sylvian fissure volume to superior sulcal space volume. T1-weighted MRI scans were acquired from three independent cohorts: the Baltimore Longitudinal Study of Aging (n = 1,032), Johns Hopkins CSF Disorders Clinic (n = 216), and the Placebo-controlled Efficacy of iNPH Shunting trial (n = 94). Preprocessing included N4 bias correction, standard-space registration, and alignment. A U-Net segmentation model with an EfficientNet-B0 encoder delineated Sylvian fissures and sulcal compartments. DESI was computed from 3D reconstructions and evaluated using Dice similarity coefficients, landmark detection error, and classification performance across diagnostic groups.

**Results:** Segmentation achieved ∼0.80 DICE for Sylvian fissures; ∼0.74 for superior sulci. On external validation, DESI discriminated NPH with DESH from non-DESH NPH with AUC = 0.99, accuracy 97.9%, and from all other groups (healthy controls, Alzheimer’s disease, vascular dementia) with perfect classification (AUC = 1.00, 100% sensitivity and specificity at threshold = 4.83). In cognitively normal adults, DESI showed only a modest age-related increase and no sex differences, confirming specificity for hydrocephalic pathology.

**Conclusion:** DESI is a robust, fully automated MRI biomarker that sensitively detects CSF redistribution characteristic of DESH in iNPH. By outperforming traditional indices and achieving near-perfect diagnostic accuracy across independent cohorts, DESI offers a noninvasive tool for early detection, patient selection for shunting, and longitudinal monitoring of disease progression.

## Introduction

Idiopathic normal pressure hydrocephalus (iNPH) is a potentially reversible cause of dementia in older adults, classically defined by the Hakim triad of gait disturbance, urinary incontinence, and cognitive decline. In its early stages, however, iNPH often manifests solely as gait impairment. Because this triad occurs frequently in the over-65 population, in neurodegenerative syndromes, prostatic urinary dysfunction, and age-related cognitive changes, clinical presentation alone is not sufficiently specific, and ancillary testing is required for reliable diagnosis1,2. Current confirmatory procedures, such as high-volume lumbar puncture and continuous lumbar drainage, can improve diagnostic accuracy (sensitivity 50-100%, specificity 60-100%) but are invasive and carry risks, headache, pain, infection, and catheter complications, that pose particular hazards for frail elderly patients3. Despite the demonstrated efficacy of cerebrospinal fluid shunting, iNPH remains markedly underdiagnosed. A recent prospective, population-based study reported a prevalence of 3.7% in individuals aged 65 and older, rising to 8.9% in those over 80, yet only a small minority of affected patients ever receive the appropriate intervention4,5.

Quantitative MRI biomarkers offer non-invasive alternatives. The Evans index, a simple linear measure defined as the ratio of maximal frontal horn ventricular width to the internal skull diameter, with values ≥0.30 indicating pathologic ventriculomegaly, has long served as a screening tool2. The callosal angle, measured on coronal slices through the posterior commissure, further refines specificity: angles <90° distinguish iNPH from Alzheimer’s disease and normal aging with ≥93% accuracy (sensitivity 97%, specificity 88%)6. Beyond these linear metrics, disproportionately enlarged subarachnoid space hydrocephalus (DESH)7 features, including tight high-convexity sulci, focally dilated sulci, and enlarged Sylvian fissures, provide disease-specific evidence of CSF redistribution in iNPH, demonstrating strong positive predictive2.

Recent advances in artificial intelligence (AI) and deep learning have enabled rapid, automated volumetric analysis of these MRI features, reducing reliance on time-consuming manual measurements. Building on these developments, we propose the Disproportionately Elevated Sulcal Index (DESI): an AI-driven volumetric biomarker that computes the ratio of Sylvian fissure volume to superior sulcal space volume from coronal MRI slices. By operationalizing DESI into a robust, reproducible metric, the DESI aims to distinguish DESH-positive iNPH from normal aging and other neurodegenerative conditions in a fully automated, non-invasive workflow.

## Method

### Data Acquisition

T1-weighted MRI data for pretraining, training, and internal validation were drawn from the Baltimore Longitudinal Study of Aging (BLSA)8 and the Johns Hopkins Center for CSF Disorders Clinic. BLSA scans comprised: at 1.5 T, SPGR acquired on three GE Signa systems (0.94 × 0.94 × 1.5 mm voxels; flip angle 45°; TE = 5 ms; TR = 35 ms; TI = 0 ms) plus one Philips Intera 1.5 T SPGR system with identical parameters; and at 3 T, sagittal MPRAGE acquired on three Philips Achieva systems (1 × 1 × 1.2 mm voxels; flip angle 8°; TE = 3.2 ms; TR = 6.5-6.8 ms; TI = 0 ms).

Johns Hopkins CSF Clinic scans were obtained on a Siemens Verio 3 T (0.78 × 0.78 × 0.80 mm voxels; flip angle 9°; TE = 3.13 ms; TR = 1800 ms; TI = 900 ms). Additionally, for the Placebo-controlled Efficacy of iNPH Shunting (PENS)9 study we acquired multi-echo T1-weighted MPRAGE with the same protocol across all sites (scan time = 2 min 54 s; resolution = 1 × 1 × 1 mm; FOV = 240 × 240 × 180 mm; 180 slices; TR/TE/TI = 14/2.1/900 ms; flip angle = 9°). All images were deidentified by stripping private metadata, converted to NIfTI format using the dcm2niix Python package, and underwent N4 bias-field correction.

### Data preprocessing

Initially, all MRI scans were subjected to a standardized preprocessing workflow to ensure data consistency and reliability. First, N4 bias field correction was applied to reduce intensity inhomogeneities10. Next, images were rigidly registered to the MNI152 standard space, aligning them to a common anatomical reference and correcting for head orientation differences, thereby enabling inter-subject comparability. Finally, scans were reoriented to the Right-Anterior-Superior (RAS) convention and aligned along the sagittal and axial axes using AC and PC coordinates extracted via BrainSignsNet11.

### Slice extraction and annotation

From preprocessed scans, we extracted every coronal slice perpendicular to the AC-PC axis between those landmarks. For model development, two slices per subject were randomly sampled from the BLSA and Johns Hopkins Clinic cohorts for training and internal validation. In the PENS trial dataset, all available coronal slices were subjected to manual segmentation in Label Studio: the right and left Sylvian fissures were delineated, and the suprasylvian sulcal spaces were segmented using the brain surface as the outer boundary. To ensure accurate identification of DESH morphology, a board-certified neuroradiologist independently reviewed every PENS scan and annotated high-convexity tightness, Sylvian fissure enlargement, and ventriculomegaly consistent with hydrocephalus.

### Sulci Segmentation Model

We employed a U-Net12 in which the encoder was initialized with pretrained EfficientNet-B0 weights and the decoder with random initialization. Coronal slices were uniformly resized to 224×224 pixels and standardized to zero mean and unit variance before input. The network’s final 1×1 convolution produces four per-pixel logits, one for each label: left Sylvian fissure, right Sylvian fissure, and suprasylvian sulci. These logits are passed through channel-wise sigmoid activations to yield independent probability maps, which are then thresholded (p = 0.5) to generate the multi-label segmentation output.

Model training was guided by a composite loss combining the Dice similarity coefficient (DSC) and Hausdorff distance (HD), the latter encouraging precise delineation of sulcal borders. To improve generalizability, we applied on-the-fly augmentation including random rotations (±15°), scaling/cropping (80-100% of original size), coarse dropout, Gaussian blurring, and additive noise. Data were split at the subject level 80% of participants from the Baltimore Longitudinal Study of Aging (BLSA) and Johns Hopkins cohorts were used for model training, with the remaining 20% reserved for internal validation to prevent data leakage. Finally, the optimized model was evaluated on the independent, multi-center PENS trial dataset to confirm its robustness across unseen scans.

### Training and Validation Setup

Model training and processing were executed on a high-performance workstation featuring dual NVIDIA RTX 4090 GPUs, 128 GB of RAM, and an AMD Threadripper Pro WRX80 CPU. Training proceeded for up to 100 epochs, or until convergence, using the AdamW optimizer with a weight-decay coefficient of 0.02.

### Disproportionately Elevated Sulcal Index (DESI) Calculation

For DESI computation (Figure 1), all scans first underwent standardized preprocessing and were realigned to the AC-PC plane. Coronal slices perpendicular to the AC-PC line were then extracted and processed using a two-dimensional segmentation approach to delineate the Sylvian fissures and sulcal spaces. Post-processing of the reconstructed three-dimensional masks was performed to remove small mis-segmented regions, particularly in the Sylvian fissures.

**Figure 1.**
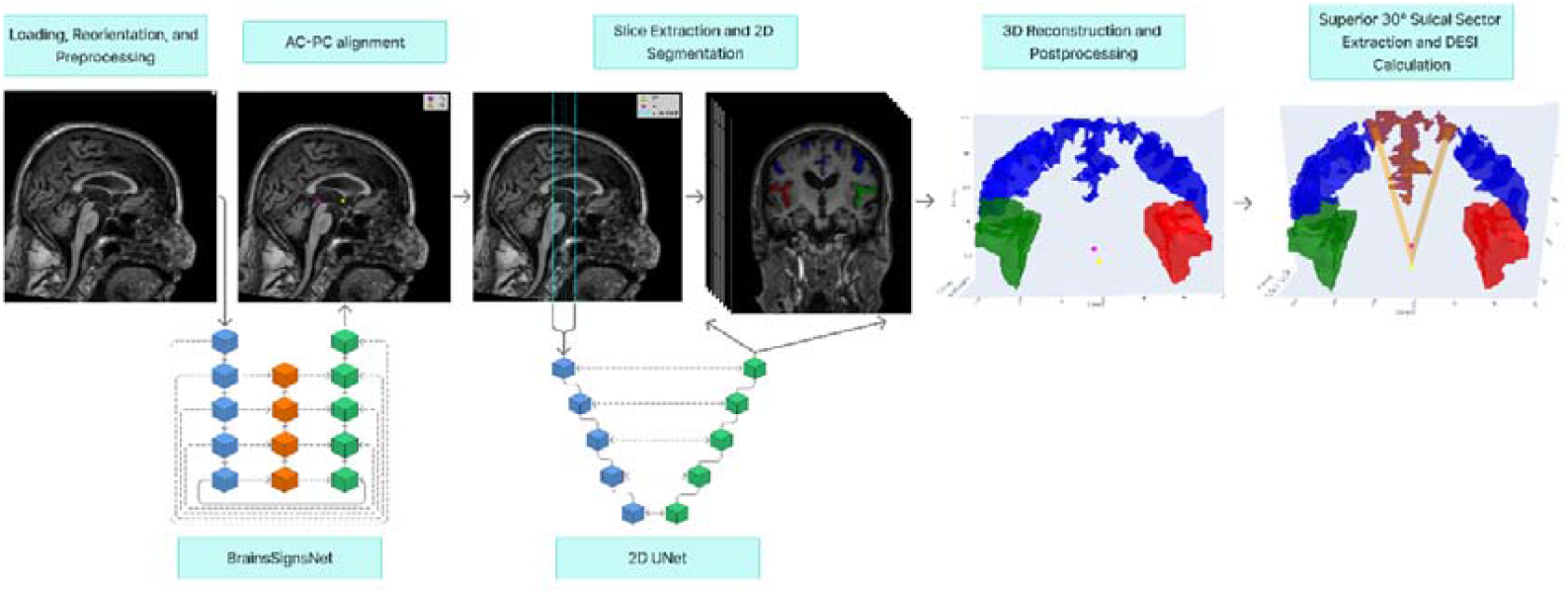
Automated Disproportionately Elevated Sulcal Index (DESI) Calculation Pipeline. AC: Anterior Commissure, PC: Posterior Commissure.

Sylvian fissure volume was calculated by summing all voxels labeled as left or right Sylvian fissure in the segmentation masks and converting voxel counts into physical volume based on the scan’s native voxel dimensions (in-plane pixel spacing and slice thickness). Superior sulci volume was quantified within a three-dimensional wedge extending superiorly from the AC-PC line with a 30° opening angle; all voxels labeled as sulcal space within this sector were included and similarly converted to volumetric units. The DESI was then defined as the ratio of Sylvian fissure volume to superior sulci volume, providing a dimensionless index of the relative enlargement of Sylvian fissures compared with superior convexity sulci.

## Statistical analysis

Continuous variables are presented as mean ± standard deviation, and categorical variables as counts (percentage). Group comparisons for normally distributed continuous data were performed using independent two-sample t-tests. Pearson’s correlation coefficient was used to assess the strength of association between pairs of continuous variables. To compare differences across more than two groups while adjusting for age and sex, we applied analysis of covariance (ANCOVA). All hypothesis tests were two-tailed, with p < 0.05 considered statistically significant.

Landmark-detection performance is reported as the mean absolute error (in millimeters) of the Euclidean distance between manually annotated and predicted commissure coordinates. Segmentation accuracy is quantified by the three-dimensional Dice similarity coefficient. Classification performance is summarized by the area under the receiver-operating characteristic curve (AUC), along with sensitivity, specificity, precision, and F_1_ score at the chosen decision threshold.

## Results Demographics

Table 1 summarizes the demographic composition of our study cohorts. The BLSA dataset comprised 1032 participants contributing 1032 coronal MRI scans (mean age 73.3 ± 13.4 years; 45.6 % male), and was used for model training, internal validation, and downstream analyses. The Johns Hopkins Clinic cohort included 216 individuals and 216 scans (mean age 72.0 ± 8.1 years; 54.1 % male), serving both training and internal validation purposes. Finally, the PENS trial enrolled 94 participants who underwent 94 scans (mean age 71.1 ± 5.5 years; 54.1 % male), which were reserved for external validation and comparative analyses. This distribution of age and sex across cohorts ensured that our model was trained and tested on diverse, multi-center data spanning a wide adult age range.

**Table 1.** Overview of participant demographics for the training and validation cohorts. “Age” denotes each subject’s age at MRI acquisition. The table also reports the gender breakdown, number of scans, total participants per dataset, and the Evans Index (presented as mean ± SD).

| Cohort | Individuals | Number of Scans | Age | Sex | Evans Index | Utilization |
| --- | --- | --- | --- | --- | --- | --- |
| BLSA<sup>8</sup> | 1,032 | 1,032 | 73.32 (13.36) | Male = 471 (45.63%)<br>Female = 561 (54.36%) | $0.28 \pm 0.03$ | Training, Internal validation, and analysis |
| Johns Hopkins Clinic | 216 | 216 | 71.99 (8.09) | F: 99 (45.8%)<br>M: 117 (54.1%) | $0.37 \pm 0.05$ | Training and internal validation |
| PENS<sup>9</sup> | 94 | 94 | 71.11(5.5) | F: 47 (50%)<br>M: 47 (50%) | $0.39 \pm 0.05$ | External Validation and analysis |

### AI Performance

On the external validation data, BrainSignsNET localized the anterior and posterior commissures with high precision, achieving median Euclidean errors below 2 mm for both landmarks. Pitch alignment was also accurate, with a mean absolute error of 1.57° and a negligible systematic bias of −0.24°.

For the multi-label sulcal segmentation, the U-Net with an EfficientNet-B0 encoder achieved the following performance: segmentation of the left Sylvian fissure yielded a mean Dice of 0.80 ± 0.15 and a mean Hausdorff distance of 6.71 ± 5.99 mm, the right Sylvian fissure achieved a mean Dice of 0.79 ± 0.16 with a mean Hausdorff distance of 7.17 ± 6.46 mm, and the superior sulcal space segmentation demonstrated a mean Dice of 0.74 ± 0.18 with a mean Hausdorff distance of 10.07 ± 9.05 mm.

### Evaluation of DESI in NPH and aging cohorts

Among cognitively normal adults without neurodegenerative disease, DESI exhibited a modest but statistically significant positive relationship with age (Pearson’s r = 0.178, p < 0.001), whereas no significant difference in DESI was observed between males and females (Mann-Whitney U = 16 697; p = 0.053) (Figure 2a-b).

**Figure 2.**
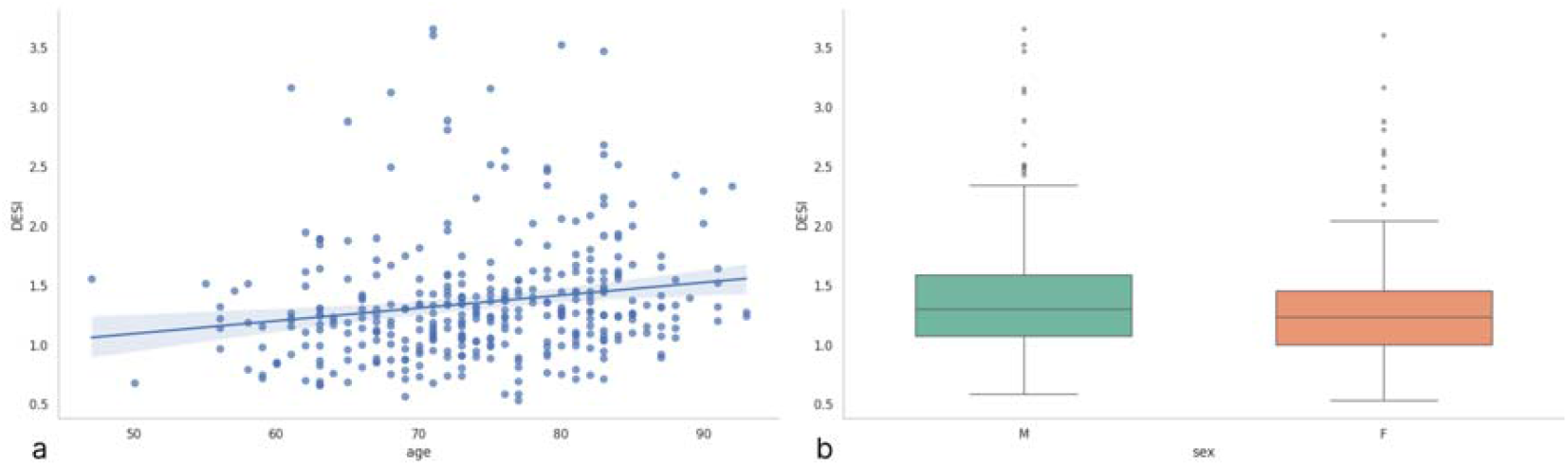
Association of DESI index with demographic factors: (a) scatter plot of age versus DESI index and (b) box plot of DESI index by sex.

In the BLSA cohort, age- and sex-adjusted ANCOVA revealed no significant DESI differences across cognitively normal, possible and probable Alzheimer’s disease, and vascular dementia groups (F (3,342) = 0.71; p = 0.546) (Figure 3a). In contrast, Mann-Whitney tests showed highly significant elevations of DESI in NPH patients with DESH versus NPH without DESH, normal controls, and dementia patients, with all comparisons reaching p < 0.0001 (Figure 3b). (Supplementary Figure 1)

**Figure 3.**
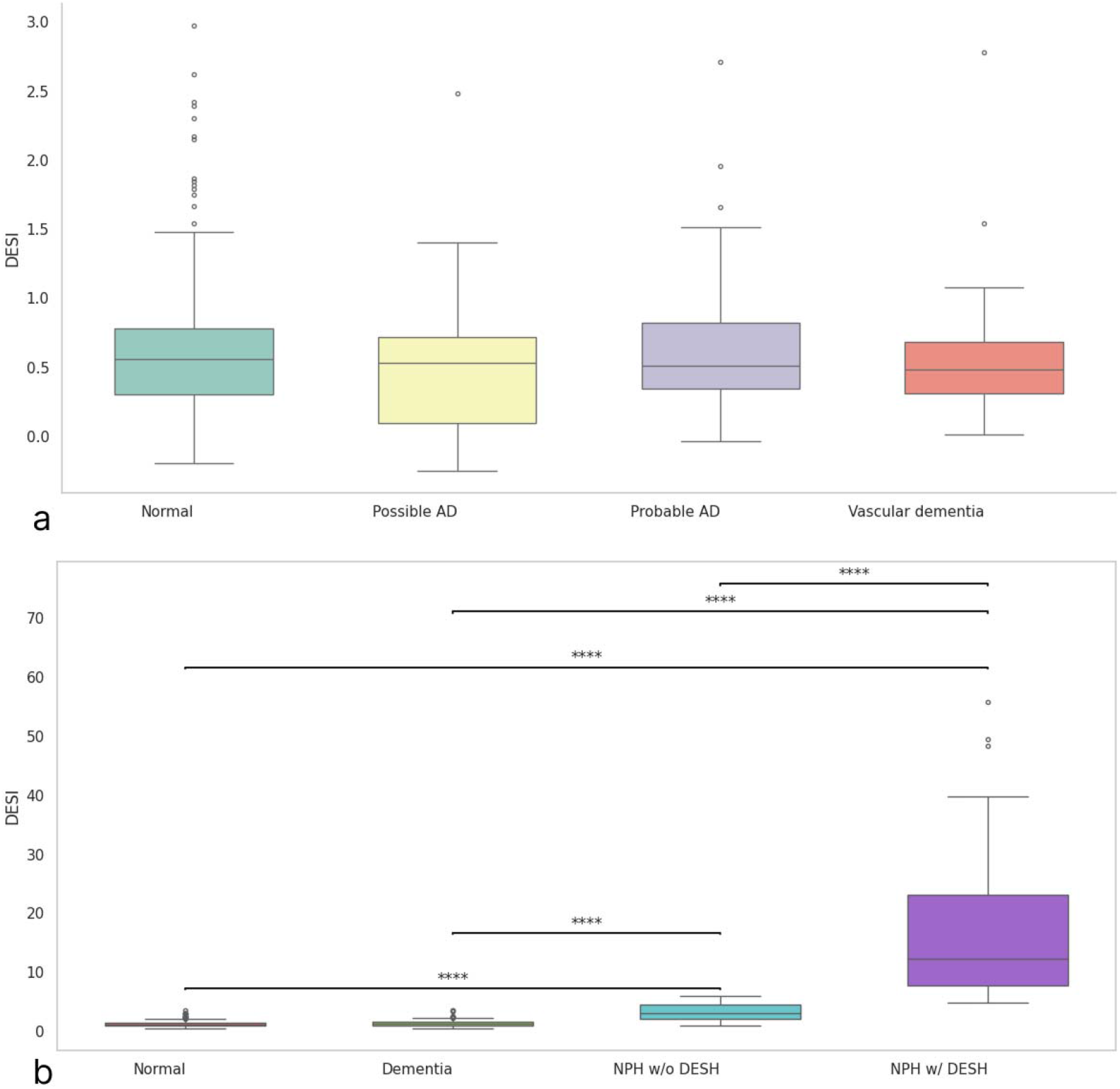
Box plots of DESI index differences across diagnostic groups: (a) normal controls versus possible Alzheimer’s disease, probable Alzheimer’s disease, and vascular dementia; (b) cognitively normal participants, participants with dementia, NPH patients with DESH, and NPH patients without DESH. **** indicates p < 0.0001.

### Classification performance

As shown in Table 2, the DESI index achieves accurate discrimination of DESH-associated NPH from both non-DESH NPH and other diagnostic groups. When separating NPH with DESH from NPH without DESH, the index attains an AUC of 0.991 with an optimal threshold of 6.10, yielding 97.9 % accuracy (95.7 % sensitivity, 100 % specificity, only 2 false negatives among 46 DESH cases). Notably, at a common threshold of 4.83, DESI perfectly distinguishes NPH with DESH from healthy controls, vascular dementia, possible Alzheimer’s disease, and probable Alzheimer’s disease cohorts (AUC = 1.00 for each; 100 % accuracy, sensitivity, specificity, precision, and F_1_ score; zero misclassifications). In a broader “NPH versus all others” analysis, the index retains excellent performance (AUC = 0.965; threshold = 1.92), achieving 90.0 % accuracy, 92.6 % sensitivity, 89.4 % specificity, and an F_1_ score of 0.798. Finally, when non-DESH NPH cases are removed from the comparator group, DESI again reaches flawless classification (AUC = 1.00; all metrics = 1.00) at the 4.83 cutoff. Together, these results underscore the high sensitivity, specificity, and overall robustness of the DESI index for identifying disproportionate sulcal enlargement in NPH and for distinguishing it from other neurodegenerative conditions.

**Table 2.** Classification performance of the DESI index for distinguishing NPH with DESH from other diagnostic groups. Class A and Class B denote the positive (NPH w/ DESH or NPH) and negative comparator groups, respectively. “Others” comprises all non-NPH subjects pooled from cognitively normal controls and participants with vascular dementia, possible Alzheimer’s disease, or probable Alzheimer’s disease. Reported metrics include area under the ROC curve (AUC), optimal decision threshold, accuracy, sensitivity, specificity, precision, and the confusion matrix for each binary comparison.

| Class A | Class B | AUC | Best Threshold | Accuracy | Sensitivity | Specificity | Precision |
| --- | --- | --- | --- | --- | --- | --- | --- |
| NPH w/ DESH | NPH w/o DESH | 0.99 | 6.1 | 0.97 | 0.95 | 1 | 1 |
| NPH w/ DESH | Normal | 1 | 4.83 | 1 | 1 | 1 | 1 |
| NPH w/ DESH | Vascular dementia | 1 | 4.83 | 1 | 1 | 1 | 1 |
| NPH w/ DESH | Possible AD | 1 | 4.83 | 1 | 1 | 1 | 1 |
| NPH w/ DESH | Probable AD | 1 | 4.83 | 1 | 1 | 1 | 1 |
| NPH w/ DESH | Others | 1 | 4.83 | 1 | 1 | 1 | 1 |
| NPH Total | Others | 0.96 | 1.92 | 0.9 | 0.92 | 0.89 | 0.7 |

Table 3 summarizes the performance of existing indexes and classification methods for detecting DESH in MRI scans. All previously proposed indexes (DESH index, Venthi index, Sylhi index) and the classification approach by Gunter et al. demonstrated high discriminatory ability between DESH and non-DESH groups, with AUC-ROC values ranging from 0.99 to 1.00. Our proposed method, DESI, achieved perfect classification (AUC-ROC = 1.00) for distinguishing DESH from non-DESH scans and from other neurodegenerative disorders, and maintained excellent performance (AUC-ROC = 0.99) when differentiating DESH from non-DESH cases with hydrocephalus. These findings highlight DESI as a robust and accurate method for automated detection of DESH across clinically relevant comparison groups.

**Table 3.**
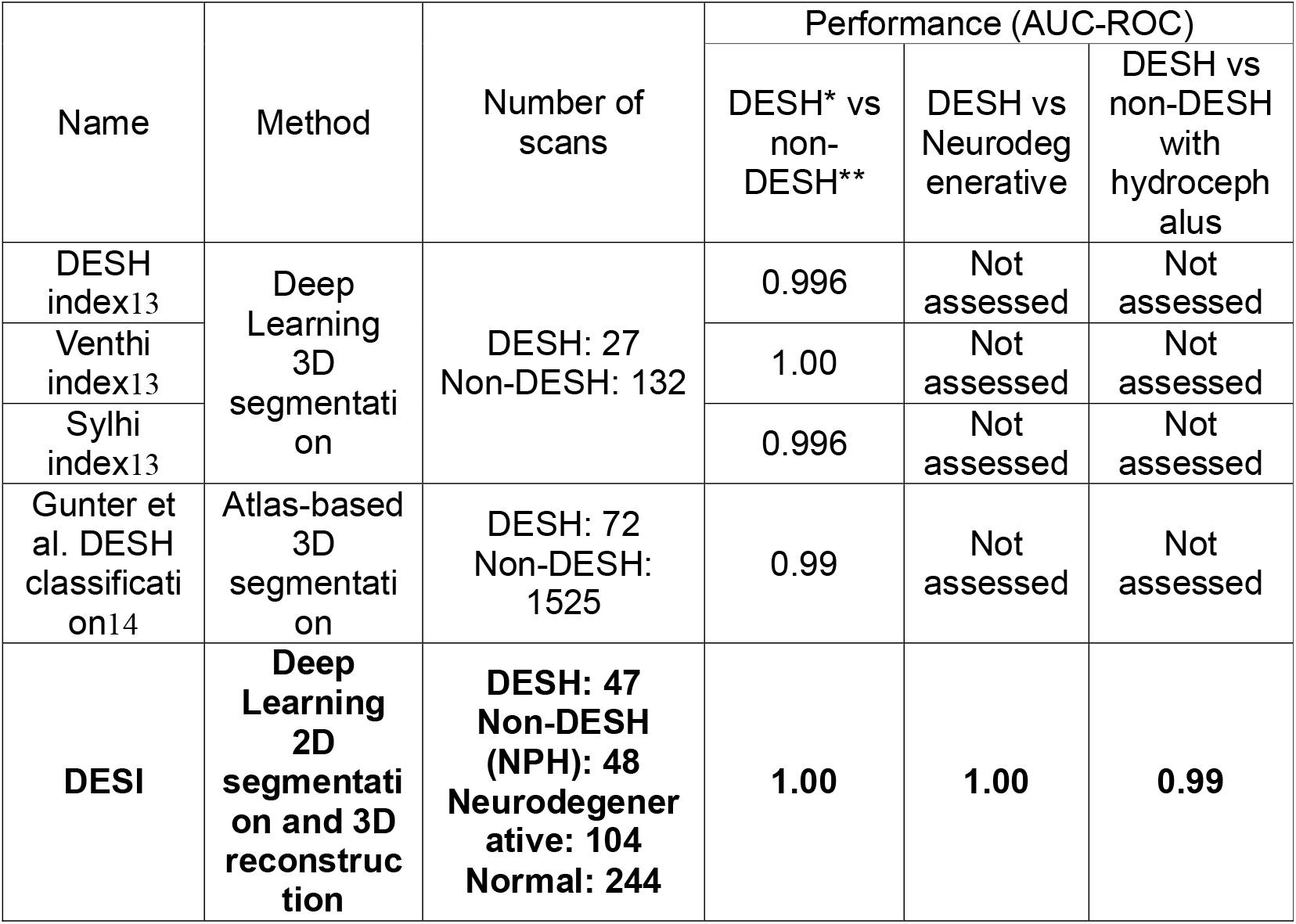
Performance comparison of existing indexes and methods for detecting DESH in MRI scans. *DESH refers to individuals with disproportionately enlarged subarachnoid spaces, whereas *non-DESH includes scans without such evidence, encompassing healthy individuals, normal aging, and other neurodegenerative disorders.

## Discussion

In this work, we introduce the Disproportionately Elevated Sulcal Index (DESI) as a novel, AI-driven biomarker for quantifying sulcal and subarachnoid space alterations in normal pressure hydrocephalus (NPH). Leveraging BrainSignsNET model for precise AC-PC alignment and a U-Net segmentation framework, we achieved robust multiclass delineation of Sylvian fissures and overlying sulcal compartments.

Applying DESI across two independent cohorts (BLSA and PENS), we demonstrated its excellent discriminatory power: AUCs ≥0.99 for distinguishing NPH with DESH from non-DESH NPH and other neurodegenerative conditions, and normal scans. In cognitively normal adults, DESI increased modestly with age but did not differ by sex, confirming its specificity for hydrocephalic pathology rather than generalized atrophy. These results establish DESI as a sensitive, reproducible metric for detecting disproportionate sulcal enlargement and highlight its potential utility for noninvasive NPH diagnosis, patient stratification, and monitoring of disease progression or treatment response.

In idiopathic normal-pressure hydrocephalus (iNPH), morphologic change extends beyond ventriculomegaly. Redistribution of cerebrospinal fluid (CSF) within the subarachnoid spaces yields a characteristic pattern, narrowing of the high-convexity sulci with disproportionate enlargement of the Sylvian fissures, that is not fully captured by the Evans Index, necessitating complementary metrics. This disproportionate CSF distribution is considered characteristic of iNPH, and several imaging indexes have been proposed to quantify it and to distinguish iNPH from neurodegenerative disorders and generalized atrophy. For example, the callosal angle (CA), which reflects upward expansion of the lateral ventricles, shows no significant change in neurodegenerative diseases other than iNPH 15. Likewise, the DESH pattern utilizes these features to aid differentiation of iNPH from other etiologies 16,17. Consistent with these observations, and as illustrated in Figure 3, our proposed index captures this iNPH-specific morphology and shows no meaningful variation with normal aging or with Alzheimer disease, Parkinson disease, or vascular dementia.

Disproportionately enlarged subarachnoid-space hydrocephalus (DESH) features often evolve gradually; consequently, binary classifications can miss early or mild presentations in which CSF redistribution has begun but not significant 18.

Continuous volumetric biomarkers address this limitation. The SILVER index 13,19, the ratio of Sylvian fissure area to vertex sulcal area, and the DESH index 13, the combined volume of the lateral ventricles, Sylvian fissures, and basal cisterns divided by the high-convexity subarachnoid-space volume, translate complex patterns of CSF shift and sulcal remodeling into unitless metrics. By providing reproducible, rater-independent measurements that sensitively track dynamic changes across compartments, these indices facilitate longitudinal disease staging, enable patient-specific risk stratification, and may inform the timing of therapeutic interventions, thereby improving diagnostic precision in iNPH.

Automated iNPH phenotyping has evolved from atlas-based and binary classifiers to deep learning-driven volumetric indices, each with distinct methodological features. Atlas-based methods14,20 harness CSF volumes from parcellated brain regions to detect tight high-convexity and DESH with high AUCs, but their reliance on precise atlas registration makes them susceptible to segmentation errors in scans with marked ventriculomegaly or other abnormalities. Another study employed fully volumetric 3D U-Net segmentation of ventricles, high-convexity subarachnoid spaces, and Sylvian regions on T1- and T2-weighted MRI, combined with a multimodal CNN classifier (regional Dice 0.60-0.85; AUC > 0.97)13. Although this framework demonstrates strong performance, it was trained on only 180 subjects and evaluated primarily against healthy controls, leaving its generalizability to other NPH mimics untested. Unlike methods that sample superior sulcal anatomy at a fixed 3 cm from the midline, our pipeline extracts coronal slices within a 30° wedge from the AC-PC axis to normalize for individual brain size differences. Applying a 2D U-Net with an EfficientNet-B0 encoder to these AC-PC-aligned slices, we derive the DESI as the ratio of Sylvian fissure to superior sulcal volumes. Validated across more than 3000 scans from independent cohorts, DESI achieves AUCs ≥0.99 for distinguishing NPH with DESH from both non-DESH NPH and other diagnostic groups, while providing a continuous measure ideal for tracking disease progression.

This study has several important limitations. First, our diagnostic groups were drawn from distinct cohorts (BLSA and PENS) acquired on different scanner platforms and protocols; despite harmonization steps (N4 bias correction, intensity normalization, AC-PC realignment), residual batch effects may have contributed to group separation. Second, manual DESH annotations and threshold selection were performed by a single neuroradiologist, without inter-rater reliability assessment, which may have introduced observer bias. Third, our cross-sectional design precludes assessment of longitudinal DESI dynamics, limiting conclusions about early detection or monitoring of disease progression. Finally, our cohorts consist primarily of North American, research-engaged older adults with relatively few comorbidities; the generalizability of DESI to more diverse populations, different clinical settings, or patients with concurrent cerebrovascular pathology remains to be demonstrated.

Future work will focus on longitudinal evaluation of the DESI to characterize intra-individual trajectories of sulcal redistribution and ventricular expansion, elucidating how early changes in CSF compartmentalization predict clinical conversion to symptomatic iNPH. By acquiring serial MRI alongside standardized assessments of gait (e.g., Timed Up and Go), cognition (e.g., Mini-Mental State Examination), and urinary function, we aim to define DESI-based staging thresholds that correlate with symptom onset, severity, and shunt responsiveness. Integrating DESI with complementary noninvasive imaging biomarkers, such as callosal angle measurements, aqueduct CSF flow quantification via phase-contrast MRI, and diffusion tensor metrics of periventricular white matter, will enable multivariate diagnostic models that capture both morphologic and hemodynamic hallmarks of iNPH. Finally, expanding these studies across diverse populations and imaging platforms will validate the robustness of a fully automated, imaging-only diagnostic workflow, ultimately facilitating earlier identification and personalized management of iNPH without the need for invasive confirmatory tests.

In conclusion, the Disproportionately Elevated Sulcal Index (DESI) represents a novel, fully automated, and noninvasive volumetric biomarker that captures the hallmark CSF redistribution of iNPH by quantifying the ratio of Sylvian fissure to superior sulcal volumes. Across three independent cohorts, DESI demonstrated outstanding discrimination of DESH-positive NPH from non-DESH NPH and other dementias (AUCs ≥0.99), while remaining robust to normal age-related sulcal widening. By moving beyond linear ventricular measures and binary DESH classification, DESI provides a continuous metric for staging disease severity, guiding patient selection for shunting, and enabling longitudinal monitoring without invasive tests. Future validation in larger, diverse, and longitudinal cohorts, as well as integration with complementary imaging features, will further establish DESI as a cornerstone of noninvasive iNPH diagnosis and management.

## Data Availability

The complete codebase for Automated DESI Measurement and detection of DESH in T1 MPRAGE scans, including all scripts and implementations of the BrainSignsNet, 2D Segmentation, and DESI calculation codes, is publicly available at https://github.com/SiavashShirzad/Disproportionately-Elevated-Sulcal-Index-DESI-. The pretrained model weights used in this study will be provided upon reasonable request from the corresponding author and are planned to be released publicly in a future update.

## Acknowledgments

This research was supported in part by the Intramural Research Program of the National Institutes of Health (NIH). The contributions of the NIH authors are considered Works of the United States Government. The findings and conclusions presented in this paper are those of the authors and do not necessarily reflect the views of the NIH or the U.S. Department of Health and Human Services. Additional funding was provided by the National Institute on Aging (U19-AG033655; P30-AG066507) and the National Institute of Neurological Disorders and Stroke (U01-NS12276). We gratefully acknowledge the PENS Trial investigators and participants for providing pre- and post-shunt surgery imaging data that served as the ground-truth validation set for this study. The PENS Trial is supported by the National Institute of Neurological Disorders and Stroke (U01NS122764) and by the Trial Innovation Network (U24TR001597, U24TR004440), funded by the National Institutes of Health.

## Notes

### Competing Interest Statement

The authors have declared no competing interest.

### Author Declarations

IRB of Johns Hopkins University gave ethical approval for this work

