## Supplementary Figure1 for "Disproportionately Elevated Sulcal Index (DESI): An automatically driven index representing disproportionate subarachnoid space enlargement in brain MRI scans"

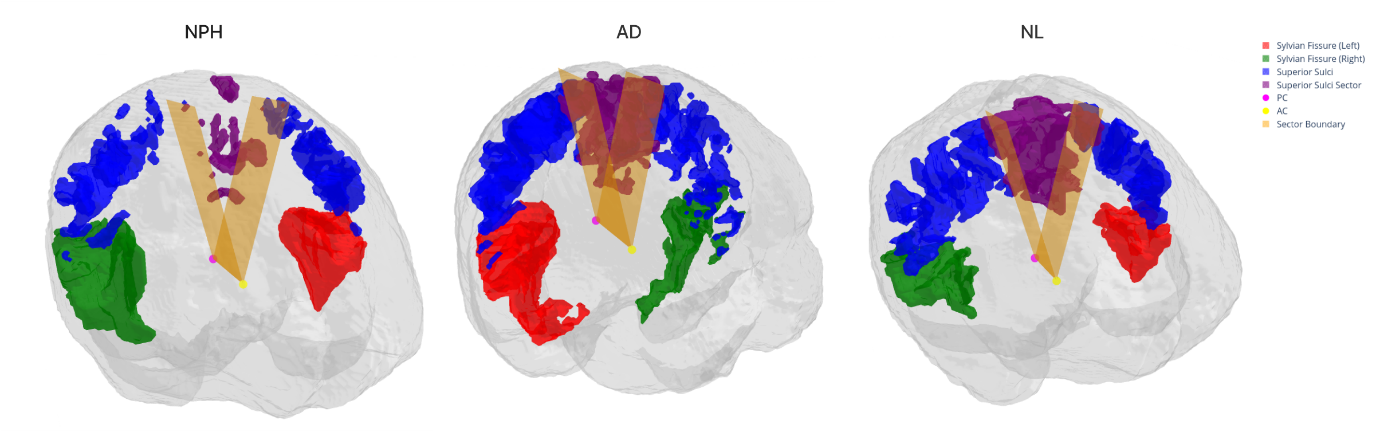

Supplementary Figure 1. Three-dimensional renderings of segmented Sylvian fissures and suprasylvian sulcal spaces within the 30° sector defined by the AC-PC line, shown for a cognitively normal individual, a patient with Alzheimer’s disease, and a NPH patient with DESH.
